# Population nuclear mitochondrial disease risk estimated from nuclear disease gene variants in a healthy older cohort

**DOI:** 10.64898/2026.05.14.26353160

**Authors:** Eloise C. Watson, Shyamsundar Ravishankar, Matthew Hobbs, Joseph Copty, Chenlong Yu, Sarah Kummerfeld, Christina Liang, Paul Lacaze, Ryan L. Davis, Carolyn M. Sue

**Affiliations:** Department of Neurogenetics, Kolling Institute, University of Sydney and Northern Sydney Local Health District, St Leonards, NSW, Australia; School of Medical Science, Faculty of Medicine and Health, Northern (Arabanoo) Precinct, University of Sydney, Camperdown, NSW, Australia; Department of Neurology, Royal North Shore Hospital, Northern Sydney Local Health District, St Leonards, NSW, Australia; Garvan Institute of Medical Research, Darlinghurst, NSW, Australia; School of Public Health and Preventive Medicine, Monash University, VIC, Australia

**Author notes:** School of Clinical Medicine, University of New South Wales, Randwick, NSW, Australia. Contributed equally (Ryan L. Davis, Carolyn M. Sue). **Correspondence:** Ryan Davis, School of Medical Science, Faculty of Medicine and Healthy, University of Sydney, Camperdown, NSW, Australia.

## Abstract

Mitochondrial diseases (MDs) are genetically and phenotypically diverse and can be difficult to diagnose. Prevalence estimates derive largely from diagnosed cases and may underestimate population MD risk. Population-based studies are limited in scope and number but indicate MD variants are common. As genomic sequencing advances have made comprehensive population-based evaluation feasible, we sought to evaluate nuclear MD variation in a population cohort to understand variant prevalence and differences in MD risk estimates

We identified disease-associated nuclear gene variants in 270 nuclear MD genes across 2,845 healthy older individuals in the Medical Genome Reference Bank. From Pathogenic or Likely Pathogenic Variants (PLPVs) we estimated autosomal recessive (AR) and autosomal dominant (AD) MD risk for individual genes and all nuclear variant-associated MDs.

We identified 554 PLPV alleles representing 357 unique variants in 145 genes. Combined AR MD risk was estimated at 25.8 per 100,000 (95% CI 18.7 to 32.9), or 1 in 3,880 individuals. *SPG7* (12.65 per 100,000; 95% CI 7.52-20.6) and *POLG* (4.23 per 100,000; 95% CI 2.10-8.01) contributed the greatest single gene AR MD risks and *OPA1* variants posed the greatest AD MD risk.

We observed a high rate of MD-associated nuclear gene variation in this healthy older cohort. The estimated lifetime AR MD risk was higher than commonly quoted prevalence estimates for *all* MDs, and the presence of common AD variants suggests variant penetrance may be lower than previously understood. These data help contextualise population MD risk and may inform clinical counselling and care.

## Introduction

Mitochondrial diseases (MDs) are individually rare but collectively considered the most common form of inherited metabolic disease.[1] However, comprehensive and accurate MD epidemiological data are challenging to obtain. MDs are defined by mitochondrial dysfunction, due primarily to defects in oxidative phosphorylation but also other essential mitochondrial functions.[2, 3] MDs are caused by pathogenic variants in either the nuclear (nDNA) or mitochondrial (mtDNA) genomes;[1] with pathogenic variants in more than 350 nuclear genes and nearly all 37 mtDNA genes implicated in disease.[4, 5] MDs can be difficult to identify clinically due to their extensive phenotypic diversity, with a disease spectrum ranging from mild single organ dysfunction to life-threatening multi-organ failure. Furthermore, MDs overlap with common clinical conditions, including diabetes, migraine and stroke.[6, 7] Diagnosis is similarly challenging, and often protracted,[8] with clinical-biochemical-histologic criteria biased toward recognised disease syndromes,[5, 9] as well as regional variability in clinical expertise and biochemical methods.[10] This means that clinical ascertainment – and epidemiological data based on clinical identification – represents a minimum detection estimate at best. In turn, this may impact planning and provisioning of health care resource.[11, 12]

Historically, MDs have been defined biochemically. The first causative nDNA variant was identified in 1995,[13] and for around two decades the available technology limited nuclear gene sequencing. In the last decade, rapid advances in sequencing technology have resulted in a dramatic and continuing increase in identification of nDNA genes and variants associated with MDs. [14] This offers new insights into mitochondrial function and MD pathophysiology,[4, 15] while also challenging the definition and diagnosis of MD. Defective oxidative phosphorylation, considered the hallmark of MDs in the biochemical diagnostic era, is less categorical in the new genetics-first paradigm of MD diagnosis,[4, 6, 16–20] where gene-product localisation, function, biochemical consequences and clinical phenotype all influence MD-gene classification.[4] Consequently, although many sources agree on a core causative gene set, there is substantial variability in the inclusion of additional genes [14, 19, 21–25], including regional versions of consensus resources, such as PanelApp (https://panelapp.genomicsengland.co.uk and https://panelapp-aus.org).[25] An exact definition to guide inclusion remains elusive,[4, 5] yet consistency is critical when estimating MD prevalence and risk. Whilst diagnostic yield improves incrementally alongside advances in knowledge and technology,[18] inconsistent genetic consideration also contributes to variable identification of affected individuals and diagnostic yield, depending on the sequencing approaches available and implemented in clinical practice.[18]

Estimates of MD prevalence from clinical cohort studies vary substantially (**Table 1**), reflecting inconsistencies in epidemiological measures, definitions of MD, inclusion criteria, tissue sampled, population diversity and case ascertainment. Reports of minimum *birth prevalence* of respiratory chain disorders (a subset of disease considered as primary MDs) in children range from 3.2 to 17.1 per 100,000 births (1 in 5,848 to 1 in 31,346 births),[10, 26–30] whilst a combined minimum *birth prevalence* estimate from paediatric and adult-onset disease was 13.1 per 100,000 or 1 in 7,634 individuals.[10, 31] A *point prevalence* of 12.5 per 100,000 (or 1 in 8,000) adults with clinically manifest MDs was obtained by Gorman *et al*. from the study of a highly specialised regional diagnostic centre in the northeast of England, whilst 23 per 100,000 (1 in 4,348) individuals – adults and children – were found to carry pathogenic nDNA or mtDNA variants that placed them and subsequent generations at risk of MD.[32] More recently, a 30-year *period prevalence* of 25.1 per 100,000 (1 in 3,989) individuals – adults and children – was estimated from hospital diagnostic codes in Ontario, Canada.[33] These data represent minimum estimates, and by comparison, estimates derived from population-based studies suggest a far higher number of individuals carry MD variants with potential risk of disease; as many as 447 to 720 per 100,000 (1 in 139 to 1 in 224) individuals.[34–39] These population studies, although unaffected by case ascertainment, have historically been limited to assessing just one or a small number of mtDNA variants, due to the technical limitations and availability of DNA sequencing,[18] and included little clinical data to evaluate any associated disease manifestations.[34–38, 40] Although new sequencing technologies are being incorporated into practice, markedly increasing genetic diagnosis rates,[19, 41–45] evaluation of MD risk specifically due to nDNA disease has until recently remained under-characterised,[32, 39, 46] despite being thought to account for around three quarters of childhood-onset MDs and one quarter of adult-onset disease.[32, 47]

**Table 1:** Epidemiological estimates of MD prevalence by region, population, study period and duration, and epidemiological measure employed.

| Author; year; region | Population | Study period (duration) | Measure | Per 100,000 (95% CI) | 1 in *n* |
| --- | --- | --- | --- | --- | --- |
| Applegarth *et al.*; [26] 2000; BC Canada | Paediatric (NS) | 1988-1996 (8 years) | Birth prevalence MDs | 3.2 (NS) | 1 in 31,346 births |
| Darin *et al.*; [27] 2001; Western Sweden | Preschool <6yo | 1984-1998 (14 years) | Birth prevalence MEs | 8.9 (5.3-14.0) | 1 in 11,247 births |
|  | Paediatric <16yo |  | Point prevalence MEs | 4.7 (2.8-7.6) | 1 in 21,000 children |
| Skladal *et al.*; [10] 2003; SE Australia | Paediatric <16yo | 1987-1996 (10 years) | Birth prevalence MRCDs | 5 (4.0-6.2) | 1 in 20,000 births |
|  | Paediatric <16yo | 1991-1994 (4 years) | Birth prevalence MRCDs | 6.2 (4.5-8.4) | 1 in 16,108 births |
| Castro-Gago *et al.*; [28] 2006; NW Spain | Paediatric <19yo | 1990-1999 (10 years) | Birth prevalence MRCDs | 14.3 (9.9-20.6) | 1 in 6,993 births |
|  | Preschool <6yo |  | Birth prevalence MRCDs | 17.1 (0-39.6) | 1 in 5,848 births |
| Diogo *et al.*; [29] 2009; Portugal | Paediatric <10yo | 1989-1996 (8 years) | Birth prevalence MRCDs | 15.4 (9.8-21) | 1 in 6,494 births |
|  |  |  | Point prevalence MRCDs | 5.4 (2.3-8.5) | 1 in 18,518 children |
| Gorman *et al.*; [32] 2015; NE England | Adult >16yo | 1990-2014 (14 years) | Point prevalence | 12.5 (11.1-14.1) | 1 in 8,000 adults |
|  | Adults and Paed |  | n- or mtDNA MD or mtDNA risk | 23.3 (14.6-34.5) | 1 in 4,348 people |
|  | Adults >16yo |  | Point prevalence nDNA MD | 2.9 (2.2-3.7) | 1 in 34,483 adults |
| Buajitti *et al.*; [33] 2022; Ontario Canada | Adults and Paed | 1988-2019 (30 years) | Period prevalence MD | 25.1 (NS) | 1 in 3,989 people |

BC British Columbia, MD mitochondrial disease, ME mitochondrial encephalomyopathy, MRCD mitochondrial respiratory chain disorder, mtDNA mitochondrial deoxyribonucleic acid, nDNA nuclear deoxyribonucleic acid, NE northeast, NW northwest, NS not specified, yo years old. **Point prevalence:** proportion of affected individuals (cases) in a defined population at a specific point in time; **period prevalence:** proportion of affected individuals (new and existing cases) in a defined population over a specified time period; **birth prevalence:** number of affected individuals (cases) identified over a specified time period from birth in a defined population relative to total number of live births in that population over that period.

Improved capability and availability of sequencing technology, along with the emergence of population level genetic reference cohorts has enabled comprehensive evaluation of MD risk, including nDNA disease. One recent study evaluated known nDNA variants in 249 mitochondrial and metabolic disease-associated genes, estimating the *lifetime risk* (the potential that a specific disease or condition will develop at any point during an individual’s lifetime; similar to genetic prevalence[48]) of autosomal recessive (AR) nDNA MD at 31.1 per 100,000 (95% CI 26.7-36.3), or 1 in 3,215, individuals.[39] This *lifetime risk* estimate, considering only AR nDNA MDs, was higher than the minimum *birth prevalence* estimates for childhood-onset MDs discussed above (3.2-17.1 per 100,000) [10, 26–30] and current estimates of *all prevalent (point* or *period prevalence)* MD from clinically based cohorts (12.5-23 per 100,000).[32, 33] *Point prevalence* estimates of AR MD are disproportionately impacted by premature mortality, especially MDs with infantile onset.[27, 29] However, even a *period prevalence* estimate of all MDs – a measure less impacted by shortened lifespan – was found to fall short of the estimated *lifetime risk* associated with only AR MDs.[33] This reveals an important gap between estimates of MD risk and rates of MD diagnosis. Whilst this gap is influenced by several factors – different epidemiological measures, diverse populations studied and reduced life expectancy – and spans an evolution in the definition and diagnosis of MD, it also highlights that some affected individuals may not present or be identified in clinical practice, even where established referral pathways, diagnostic guidelines and/or regional testing laboratories exist.

As well as considering MD definition, technical capabilities and gene sets, consideration must be given to the populations studied. The aggregation of large, publicly available genomic databases [49–51] allows evaluation of nDNA (and increasingly mtDNA [38, 52, 53]) variants in the population. However, by virtue of their aggregation, these databases may not be representative population samples, may reflect varied research cohort sampling (disproportionately representing certain disease-affected individuals or their relatives) and be subject to *survivor bias*.[54] The Medical Genome Reference Bank (MGRB) is a purpose-designed biobank of unrelated, healthy, older Australians, primarily of non-Finnish European genetic background, who lived to at least 70 years of age without any history of cancer, cardiovascular disease, or dementia.[50, 54] This intentionally curated and consistently sequenced, older adult, disease-deplete biobank [50] offers a reference population to study population pathogenic variation associated with MD. As over half of all variation is observed to be unique, even in very large datasets [55], it is relevant to consider known and putative PLPVs through implementation of standardised variant classification guidelines.[56]

Careful curation of an MD gene set enables comprehensive evaluation of pathogenic variation in relevant genomic reference databases. Variant frequencies derived from such databases enable estimates of population-level MD risk associated with both specific genes and across all MD-associated genes.[39] Population risk estimates may in turn provide a denominator against which to evaluate MD clinical identification, contextualise interpretation of sequencing results, provide focussed counselling, and inform clinical decision making, referral pathways and appropriate healthcare resource planning. We sought to use this approach to estimate population risk for all nDNA MD genes across all inheritance types to better understand the divergence in MD epidemiological estimates between clinical cohort and population-based studies. To do this, we compiled a consensus MD gene set, evaluated the prevalence of pathogenic nDNA MD variants in the MGRB cohort, from which we estimated *lifetime risk* of nDNA MDs in the population, and compared to prior estimates from clinical cohorts.

## Methods

### Cohort

The MGRB cohort [50] used for this study is a consented biobank of genetically characterised and deeply phenotyped healthy older Australians of predominantly European descent drawn from two studies; the ASPREE study [57] and the 45 and Up study.[58] Whole genome sequencing (WGS) of blood DNA was performed on Illumina HiSeq X sequencers at the Garvan Institute, Sydney. Sequence data was processed following GATK best practices [59] and mapped to human reference genome build 37 (GRCh37).[50] The publicly available genetic variation data used in this study was extracted from the custom-built online Vectis platform (https://sgc.garvan.org.au/explore [54]) between January and April 2021 and contained genome sequences of 2,845 unrelated individuals at the time of access. Variants arising in curated MD gene sets (below) were extracted for analysis.

### Gene sets

MD gene lists published by six recognised adult and paediatric MD clinical-research groups, [14, 19, 21–24] were collated to generate an initial list of putative MD genes (to June 2020). For the purpose of this study, MDs are characterised by dysfunctional mitochondria stemming primarily from defects in oxidative phosphorylation or other essential mitochondrial functions.[2, 3] Genes were checked for updated mitochondrial relevance and disease association using online sources, including PubMed,[60] ClinVar,[61] OMIM,[62] and, once available, PanelApp England and PanelApp Australia,[25] to form a final comprehensive MD gene list (**Suppl1_MDgenes**). Recognising the variability in genes included by different sources, a “consensus” (high confidence) subset of MD genes was also generated; defined as genes included by at least 4 of 6 sources and confirmed as being associated with MD on manual review. In addition, for genes with clear MD association but more recently described, associated with, or categorised as MD, discretion was applied, and fewer sources were required for inclusion in the consensus subset.

### Variants

Variants of relevance were identified by individual gene searches of the MGRB Vectis platform (accessed between June 2019 and April 2021). Initial variant filtering was undertaken utilising tools embedded within the platform: variants were retained for further evaluation if they met one of three criteria: 1) a ClinVar rating of Pathogenic or Likely Pathogenic (based on embedded ClinVar; static versioning from 2018 – manually updated in November 2025)), or, for those with conflicting ClinVar classification, where weight of evidence favoured pathogenicity, 2) consensus deleterious computational prediction (based on embedded prediction tools, PolyPhen[63] and SIFT[64]), 3) functional consequence of frameshift, nonsense variant (start loss, stop gain) or splice region variant. Retained variants were then assigned an initial American College of Medical Genetics and Genomics (ACMG) classification [56] utilising the VarSome API (v9.4.4, accessed June 2021).[65] Variants assigned ‘Pathogenic’ or ‘Likely Pathogenic’ were retained for manual review. Variants with a rating of ‘Uncertain Significance’ and those where ClinVar and VarSome classifications differed were also reviewed; only those with evidence favouring pathogenicity were retained. Manual review (conducted by ECW) was performed to confirm QC parameters, update MD relevance and ClinVar rating (most recently November 2025), review and refine ACMG variant classification, and determine mode of inheritance.

### Disease risk

To estimate MD risk based on allele frequency, MD risk was calculated for each gene and summed across all genes. For AR disease, the risk (*R_i_*) for a given nDNA gene (*i*), was calculated from the square of summed pathogenic variant allele frequencies (*q_ij_*), assuming mutual independence of variants, Hardy-Weinberg equilibrium and full penetrance of biallelic mutations:

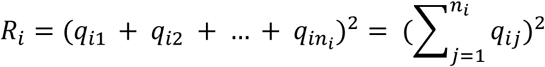

The combined risk of AR MD (*R_total_*) was calculated by summing the risk of disease associated with each gene, assuming independent occurrence of disease related to each MD gene:

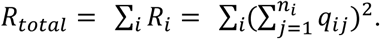

AD MD risk is more challenging to quantify, due to penetrance over the life course. For the older MGRB cohort, relevant age-specific penetrance may reasonably be assumed to approximate lifetime penetrance. For the purposes of this study, assuming rare variation, the risk (*R_i_*) for AD MD due to an individual variant (*i*), with allele frequency (*q_i_*), carrier frequency (*2q_i_*) and lifetime penetrance (*P_i_*), was estimated as:

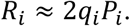

The AD MD risk per gene was calculated as the sum of AD MD risks for each variant in that gene and the combined AD MD risk (*R_total_*) for all AD MD gene variants. Assuming independent occurrence of variants and negligible variant overlap, AD MD risk was calculated by summing the risks for each variant:

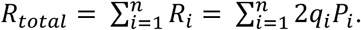

Penetrance was determined where possible from the literature or extrapolated from available data. For genes associated with both monoallelic and biallelic disease, specific variant inheritance pattern was used where available or where reasonable prediction could be made. Otherwise, the more conservative AR MD risk was used for estimates. For variants known to cause both AR and AD disease, risk calculation included both inheritance mechanisms. No pathogenic X-linked variants were identified in this cohort. ***Figure 1*** outlines the processes of variant identification, filtering, classification and review, and the subsequent process of risk estimation.

**Figure 1.**
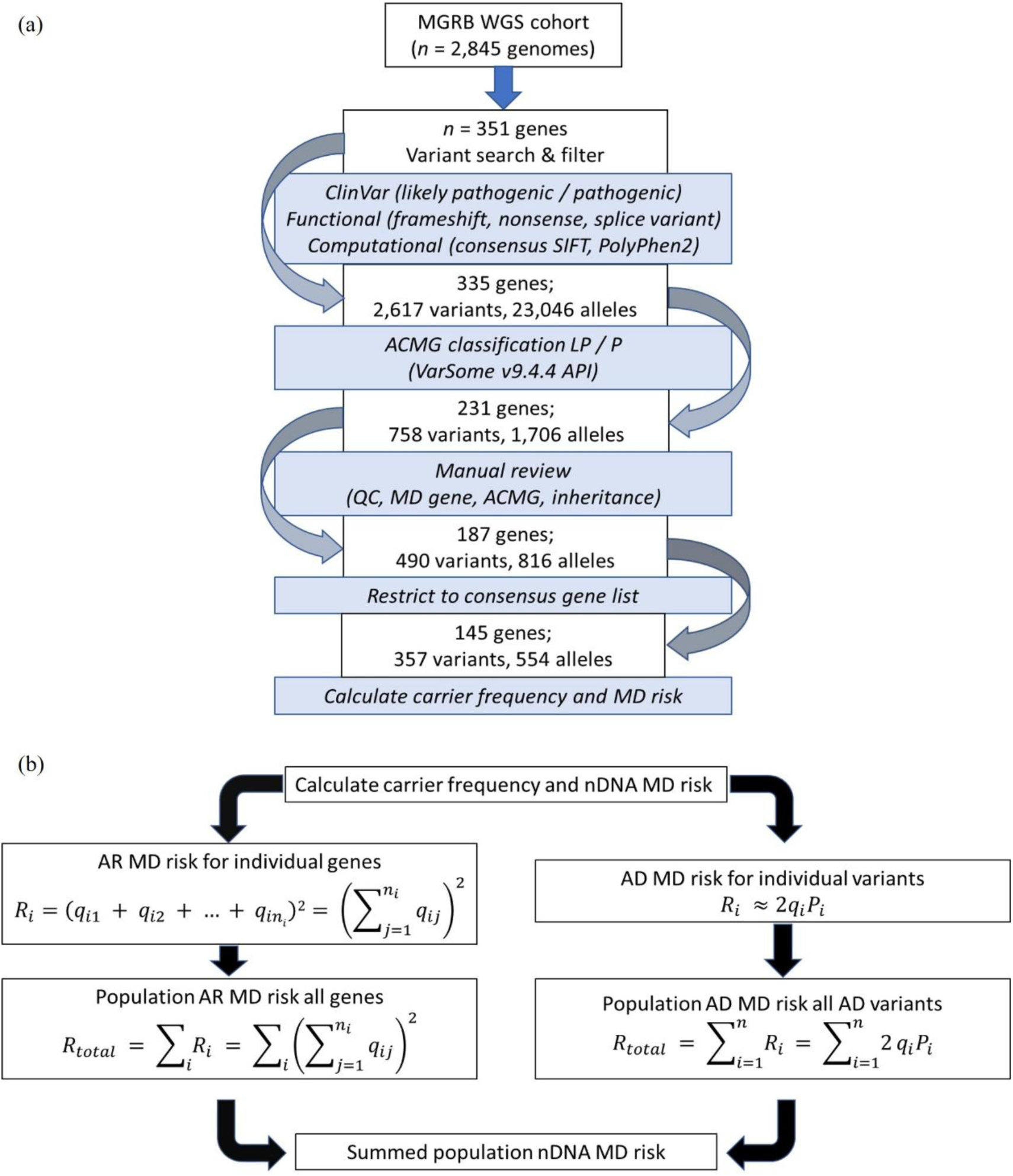
(a) Flow diagram outlines nDNA variant identification, filtering, classification, and review steps, together with counts of alleles, variants and genes retained at each step, followed by **(b)** which depicts calculation of carrier frequency and nDNA MD risk for AR and AD variants, respectively. ACMG American College of Medical Genetics and Genomics; AD autosomal dominant; AR autosomal recessive; MD mitochondrial disease; MGRB Medical Genome Reference Bank; LP likely pathogenic; P pathogenic; QC quality control; WGS whole genome sequencing.

### Analysis

Data was analysed using *R* version 4.3.1 and *RStudio* 2024.04.1+748. Individual variant risk and confidence intervals were calculated with the RStudio package PropCIs using Clopper Pearson exact intervals; for summed AR MD risk, the delta method was employed to estimate variance and confidence intervals.

## Results

### Consideration of disease gene sets

Causative MD genes from studies authored by six MD clinical-research groups [14, 19, 21–24] were compiled to generate an initial list of 422 nuclear genes considered to be associated with MD. On review, 71 of these genes did not have a clear MD association (non-mitochondrial, non-MD or no reported phenotype) and were removed, leaving 351 genes forming a “comprehensive” list. From this comprehensive list, 270 genes, occurring in 4 or more of the sources or by discretion following manual review, met criteria for a “consensus” list (**Suppl1a_MDgenes_consensus**); the difference of 81 genes were considered in the comprehensive set only (**Suppl1b_MDgenes_comprehensive)**. A small number of genes were reported in association with MD after finalisation of the gene sets but could not be included in this study; they have been included for reference in **Suppl1c_MDgenes_future**. ***Figure 2*** demonstrates the overlap with and variation from these consensus and comprehensive MD gene sets by source. Of the 270 consensus genes, 151 (56%) were included by all six sources, 57 (21%) were included by five of the six sources and another 27 (10%) were considered by four of the six sources. The remaining 35 genes (13%) occurred in three or fewer sources and were included by discretion based on manual review. The 81 additional genes in the comprehensive list included a combination of 1) 25 genes (31%) likely associated with MD but with minimal reported cases or evidence, 2) 20 genes (25%) associated with mitochondrial dysfunction but causing a phenotype atypical for MD, and 3) 36 genes (44%) associated with diseases more commonly considered in alternative categories (e.g., inborn error of metabolism (IEM), muscle disease, movement disorder).

**Figure 2.**
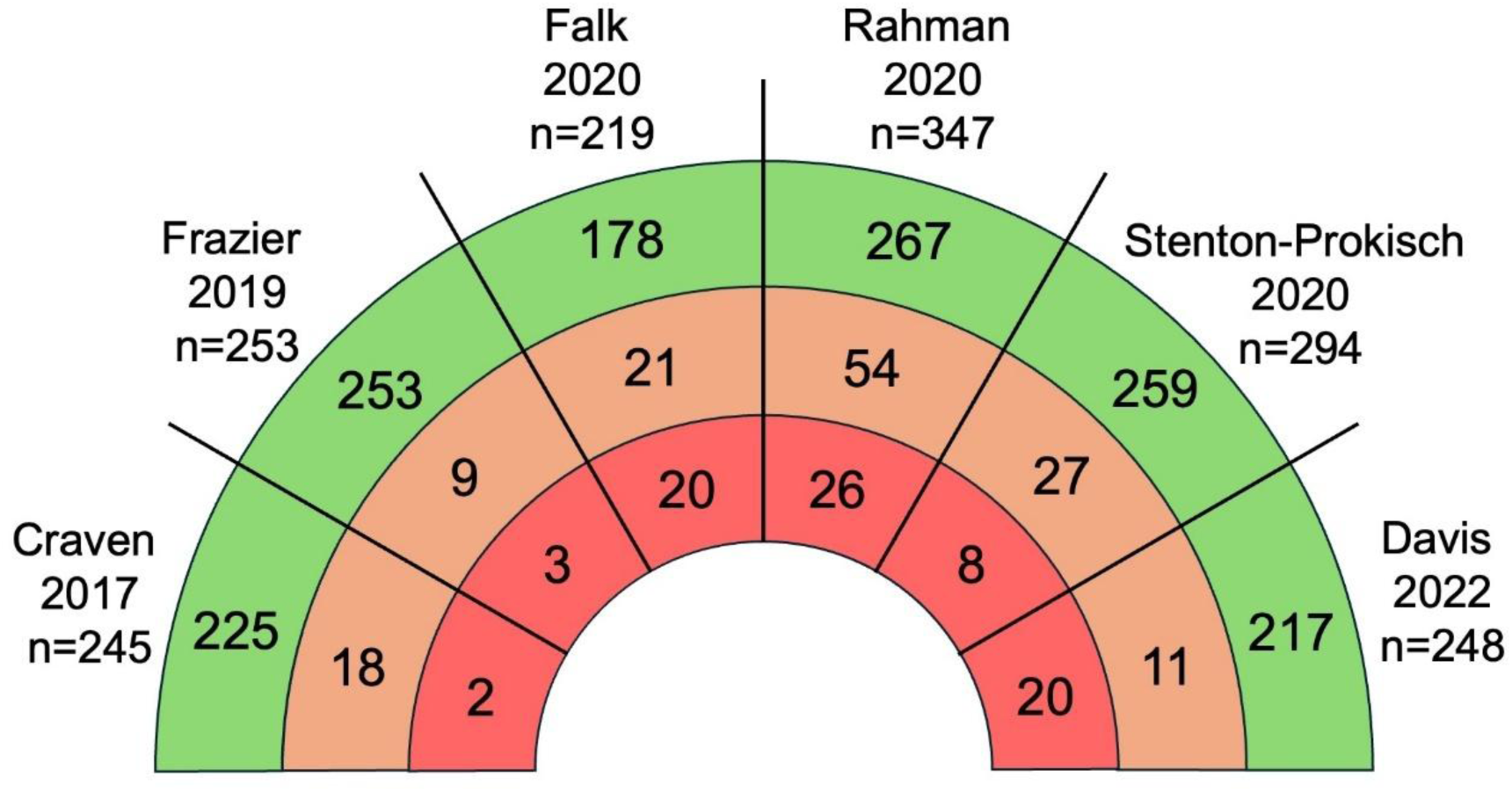
Integration of nDNA MD gene lists from the six identified MD clinical-research group published sources demonstrated variability in genes associated with MD. The figure indicates the total number of MD genes listed by each source, as well as the number of genes from each source that contributed to the final 270 consensus (green band) or additional 81 comprehensive (orange band) MD gene sets in this study. The consensus set contained *n* = 270 genes, and an additional *n* = 81 genes were included in the comprehensive set (total genes in the comprehensive set *n* = 351). The red band (containing *n* = 71 excluded genes) shows the numbers of genes from each source that were not associated with MD on review and were excluded from the final MD gene sets evaluated. Bands are not proportionate.

### Variant filtering and prioritisation

Initial extraction of variants from the comprehensive gene list retained 23,046 alleles representing 2,617 unique variants in 335 genes. Of these variants, 12,484 alleles representing 1,780 unique variants in 256 genes segregated with the consensus gene list. Following ACMG classification and manual review, a total of 816 alleles representing 490 different variants in 187 genes were retained from the comprehensive list, of which 554 alleles representing 357 unique variants were in 145 genes from the consensus list (see ***Figure 1a***). The variants meeting analysis criteria were tabulated with allele frequencies and calculated MD risks in **Suppl2_ARvariants** and **Suppl3_ARrisk**. As expected, all identified variants were heterozygous in this healthy population cohort, with only a small number of identified variants associated with monoallelic MD (*n* = 7), or biallelic and monoallelic disease (n = 2) (summarised in **Suppl4_ADvariants**, together with penetrance estimates and risk).

### Autosomal recessive MD risk

There were 549 alleles representing 352 unique variants with AR inheritance in 143 genes from the consensus gene set. The overall risk of MD related to AR nuclear variants in this cohort, implementing the consensus MD gene list, was estimated at 25.8 (95% CI 18.7 to 32.9) per 100,000, or 1 in 3,880 individuals. The genes conveying the highest risk of AR MD were *SPG7* (12.65 per 100,000; 95% CI 7.52-20.6), *POLG* (4.23; 2.10-8.01), *PPA2* (1.78; 0.73-3.93), *LIPT1* (1.00; 0.35-2.50) and *TMEM126B* (0.52; 0.15-1.52), followed by *FBXL4*, *COQ8A*, *ETFDH*, *GFM1*, *COASY*, *OPA1*, *MRPS34*, *SACS*, *NDUFB3*, *LARS2*, *SURF1*, *RMND1*, *POLRMT*, *PITRM1*, *FARS2*, and *ELAC2* (***Figure 3*** and **Suppl3_ARrisk**). Approximately 1 in 44 healthy older individuals carried a pathogenic variant in *SPG7* (64 alleles, 13 unique variants, 2.2% of individuals) conferring an estimated lifetime risk of 12.7 (95% CI 7.52-20.6) per 100,000 individuals. The next greatest risk was associated with *POLG*, where 1 in 77 individuals harboured a pathogenic variant (37 alleles, 10 unique variants, 1.3% of individuals) and the estimated lifetime risk was 4.23 (95% CI 2.10-8.01) per 100,000. The finding that *SPG7* and *POLG* contributed the highest estimated risk for AR MDs in the *MGRB* cohort is consistent with observations of the most common causes of AR nDNA disease from the northern England clinically-based adult cohort, although prevalence was only 0.8 per 100,000 and 0.3 per 100,000 respectively in that study.[32] Amongst all 270 consensus MD genes, a relevant AR MD risk allele was found in 143 (53%) genes and in one third of these (51 of 143; 36%) only a single pathogenic allele was identified.

**Figure 3.**
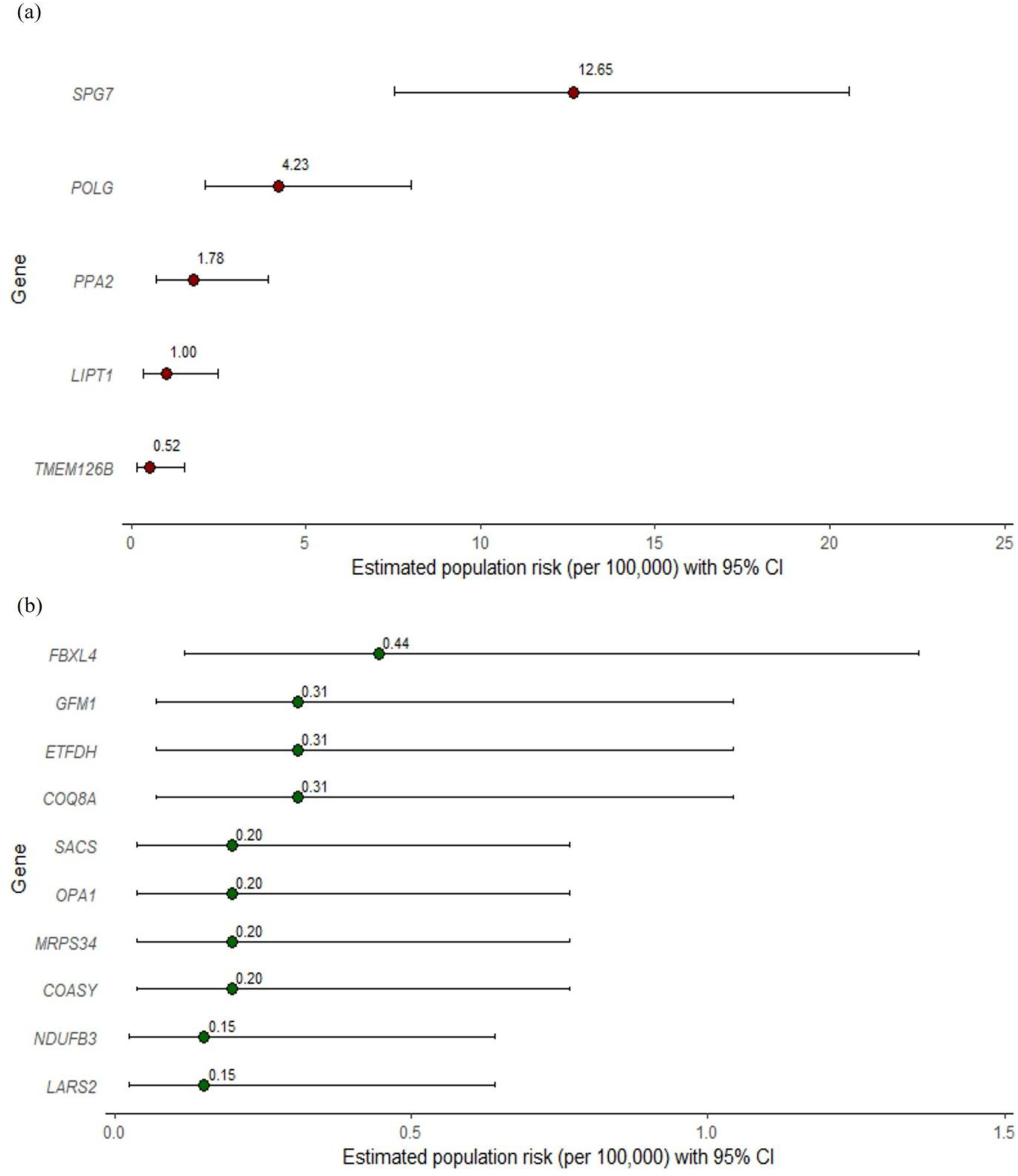
Estimated population risk of AR MD per 100,000 individuals for ***(a)*** the five highest risk genes and ***(b)*** the next 10 highest risk genes identified in this cohort. Note *x*-axis scale differs between figures.

### Variation in AR risk estimates based on gene selection

Compared to risk estimates derived from the consensus gene set, implementing the comprehensive list resulted in a considerably higher MD risk estimate of 41.8 per 100,000 (95% CI 33.1 to 50.5) or 1 in 2,392 individuals. However, genes more classically associated with metabolic disorders rather than MD (*ACADM* (6.82 per 100,000; 95% CI 3.69-12.03), *BTD* (1.12 per 100,000, 95% CI 0.40-2.71), *ACADVL* (0.89 per 100,000, 95% CI 0.30-2.28), *SLC22A5* (1.12 per 100,000, 95% CI 0.40-2.71)) and AR Parkinson’s Disease (PD) due to mitochondrial dysfunction (*PRKN* 3.57 per 100,000, 95% CI 1.72-6.96), contributed substantially to this difference, highlighting the impact of gene selection. Applying the AR gene set used by Tan *et* al.,[39] which also included a number of genes more classically associated with metabolic disease, yielded an estimate of 36.9 per 100,000 (95% CI 28.6-45.2), or 1 in 2,712 individuals in this study cohort. This is slightly higher than the reported estimate from the Tan *et al*. study based on the global gnomAD database (31.1 per 100,000, 95% CI 26.7-36.3) and from their in-house database (31.8 per 100,000, 95% CI 20.9-50.6), potentially reflecting the inclusion of novel variants alongside known variants. However, it is also less than their estimate from the European (non-Finnish) gnomAD dataset (48.4 per 100,000, 95% CI 40.3-58.5), despite the MGRB reflecting similar population ancestry, consistent with the risk-deplete nature of the cohort.[50] There were also notable differences in summed MD risk estimates when implementing ‘green’ gene lists from PanelApp England (*n* = 298 genes; 27.7 per 100,000, 95% CI 20.5-34.9, or 1 in 3,610 individuals) and PanelApp Australia (*n* = 332 genes; 39.5 per 100,000, 95% CI 30.9-48.1, or 1 in 2,532 individuals). The AR MD risk estimate based on the PanelApp England list was close to that of the consensus MD list used in this study whereas the estimate based on the PanelApp Australia list – which includes several metabolic and PD genes under the broader umbrella of MD – more closely approximated the estimate from the comprehensive list used in this study.

### Comparison of AR MD risk with prior epidemiological studies

Variant allele frequencies and MD risk from the 15 highest nDNA-MD risk genes identified in this study were compared with those from the study by Tan and colleagues, the only other similar study of nDNA MD risk.[39] Substantial overlap was evident for the highest MD risk genes (allowing for differences in genes sets) and for estimated MD risks associated with individual genes (as outlined in ***Table 2***). By comparison, nDNA MD prevalence estimates from the northeast England adult clinical cohort (2.9; 95% CI 2.2-3.7 per 100,000 individuals; of which 1.45 per 100,000 had AR MD) [32] were an order of magnitude lower than lifetime AR MD risk estimates (25.8; 95% CI 18.7-32.9 per 100,00 individuals). Birth prevalence estimates, unaffected by the premature mortality associated with MDs, may be more comparable to lifetime risk estimates. Studies reporting this measure are restricted to paediatric onset MD, and don’t consider intrauterine death.[10, 26–29] However, estimates span a wide range from 3.2 to 17.1 per 100,000 [10, 26–29] (***Table 1***), with those at the higher end of the range approaching *lifetime risk* estimates.

**Table 2:**
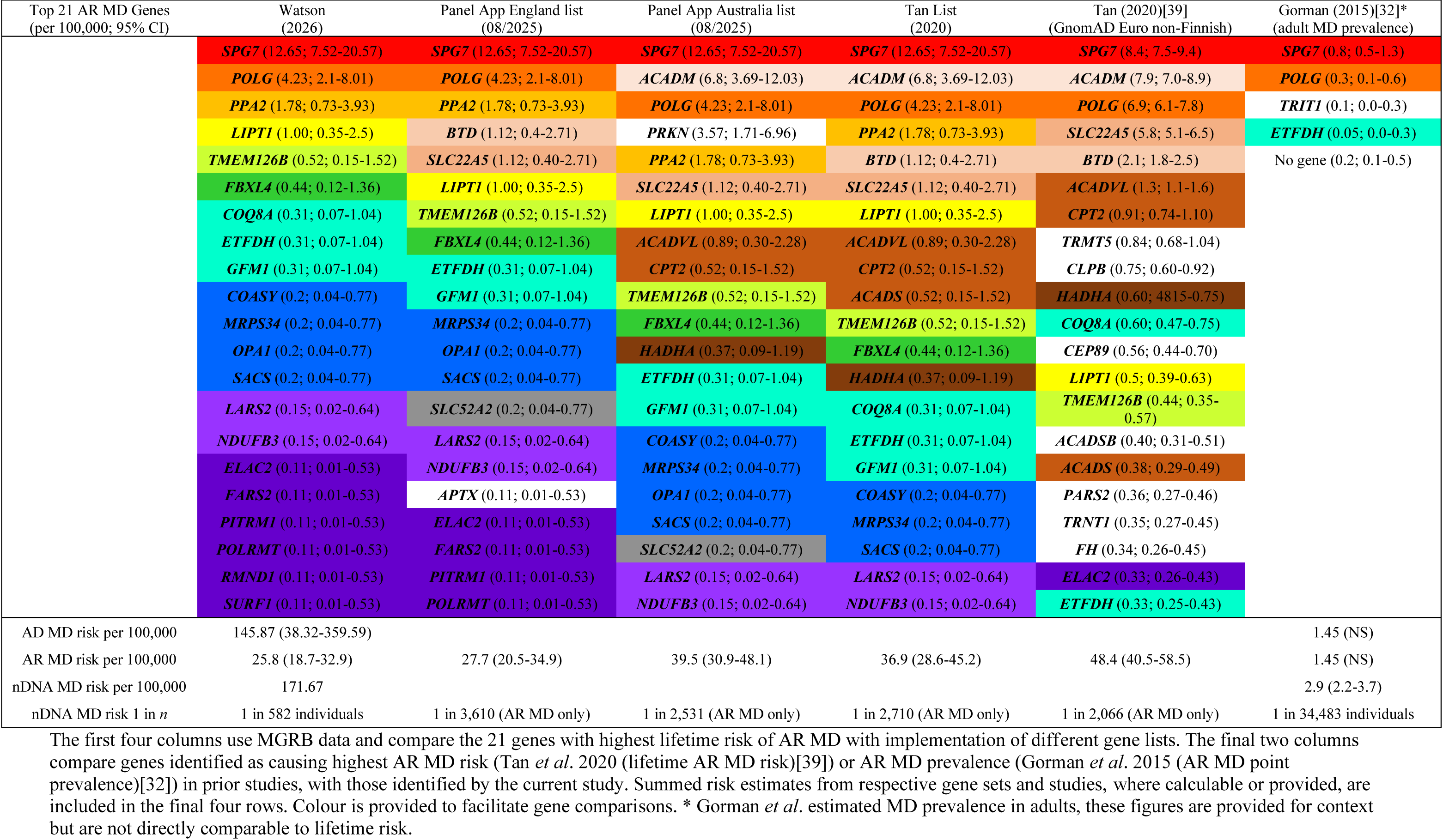
Comparison of genes with highest estimated MD risk or disease prevalence between gene sets and studies.

### Autosomal dominant MD risk

In this healthy older cohort, 11 alleles carried variants in seven different genes (*MFN2*, *OPA1*, *CYCS*, *TWNK*, *IDH2*, *SPG7* and *POLRMT*) associated with a dominant mode of inheritance and potential for monoallelic clinical manifestations. *SDHA* variants were not considered, as the disease association in monoallelic form appears exclusively with phaeochromocytoma paraganglioma syndrome 5 (PPGL5). Six of these seven genes were in the consensus MD list (*IDH2* was not), and only one variant was present with an allele count >1 (*SPG7* (NM_003119.4(*SPG7*):c.1450-1_1457del); allele count = 3), noting controversy regarding monoallelic manifestations of this variant. Of the nine consensus variants, two can potentially cause MD in mono- or biallelic state (four alleles; one in *MFN2* (NM_014874.4(*MFN2*):c.2119C>T (p.Arg707Trp)) and the aforementioned three in *SPG7*), whilst the remaining seven variants (seven alleles) associate with MD in the monoallelic state. Considering MD genes in the consensus list, as many as 1 in 316 individuals carried a variant with potential monoallelic manifestations (1 in 474 if the *SPG7* variant is not considered). Monoallelic nDNA MD risk was calculated as outlined above, incorporating an estimate of penetrance, and is summarised in **Suppl4_ADvariants**. Estimated this way, AD variants contributed additional risk for nDNA MD of 145.87 (95% CI 38.32-359.59) per 100,000 individuals, or 209.14 (95% CI 77.43-458.46) per 100,000 for the comprehensive gene set, although the small total number of AD variants limits precision. As a healthy older population assumed to be deplete of pathogenic AD alleles,[50] this represents a minimum estimate. However, it is substantially higher than encountered in clinical practice, suggesting variant penetrance may in fact be substantially lower than assumed from clinical studies alone.

### Comparison of AD MD risk with prior epidemiological studies

We found *OPA1* variants to be the most common AD MD variants in this cohort. Incorporating penetrance estimates based on prior reports,[66–70] *OPA1* variants presented the greatest population risk for AD MDs at 56.2 (95% CI 8.5-253.7) per 100,000 individuals. *OPA1* variants were also observed as a relatively common cause for AD MD in the clinical cohort-based study by Gorman *et al*.,[32] although the prevalence was estimated at 0.4 (95% CI 0.2-0.7) per 100,000 adults. Whilst *OPA1*-associated MD was observed commonly by Gorman *et al*.,[32] the most observed pathogenic AD MD gene in that study, *PEO1* (*TWNK*; prevalence 0.7 (95% CI 0.4-1.2) per 100,000 adults), had only a single pathogenic AD variant observed in the current study, with population risk estimated at 21.09 (95% CI 0.8-195.7) per 100,000 individuals. It may be that *TWNK* has higher penetrance and is more likely to cause multisystem disease, therefore being selected out of the healthy older MGRB cohort considered here. However, with the low numbers of AD variants, a relatively small cohort size and the inclusion of only healthy participants, the precision, observation, and inference is limited.

### Combined AR and AD risk

The combined lifetime risk for AR and AD nDNA MD in this healthy population cohort was estimated at 172 per 100,000 individuals, or approximately 1 in 600 individuals. However, the greatest contribution to this risk arose from AD MD, which has not previously been systematically estimated and should be interpreted with caution given the small numbers, wide confidence intervals and likely overestimate of risk through use of penetrance from clinical ascertainment, as observed in other conditions.[71–73] Overall, *OPA1* conveyed the greatest estimated single-gene risk of nDNA MD (1 in 1,779 individuals) whilst *SPG7* (1 in 7,905 individuals) and *POLG* (1 in 23,641 individuals) conveyed the greatest estimated risk of AR nDNA MD. Disease associated with all these genes is relevant to both adult and paediatric clinicians, as individuals may present throughout the lifespan.

## Discussion

Using a population-based approach in this healthy older Australian cohort, we estimated population AR nDNA MD risk to be 25.8 (95% CI 18.7-32.9) per 100,000 individuals, or 1 in 3,880 individuals. This is greater than the commonly quoted risk of 1 in 5,000 individuals for *all* (nDNA and mtDNA) MD across the lifespan,[10] and comparable to the only other AR MD risk estimate, by Tan and colleagues in 2020, which used a similar methodological approach.[39] Approximately 1 in 300 to 500 individuals carry a variant potentially associated with AD MD, and based on current penetrance data, AD MD risk was estimated at 145.87 (95% CI 38.32-359.59) per 100,000 (or 1 in 686) individuals, higher than encountered in clinical practice. Although the small numbers of variants and broad confidence intervals mandate caution in interpretation of this AD estimate, it suggests penetrance in the population context may need revision. For all nDNA MDs, genes with the highest estimated population MD risk in this study (*SPG7*, *POLG, OPA1*) accord with commonly identified pathogenic genes in clinical practice,[32] supporting the utility of this population-based approach to estimate MD risk, although absolute estimates differ greatly between prevalence (point, birth and period) and risk data. This population-based data is complementary to that derived from clinical cohorts and integrating these differing estimates can provide valuable insights for MD care. Despite various contributing factors – including epidemiological measures, premature MD mortality and advances in technological diagnostics – the findings suggest that incomplete ascertainment of MD is relevant. This may reflect a constellation of factors including primary care recognition, tertiary referral pathways, locoregional expertise, and specific diagnostic or sequencing approaches.

Lifetime risk encompasses both adult and paediatric disease, which are generally considered separately in MD as largely non-overlapping groups [10, 27, 29], justifying incorporation of estimates from both. Adult MDs are recognised as predominantly attributable to mtDNA variation,[32, 74] whilst the majority of paediatric MDs are attributable to AR nDNA disease.[10, 74–77] Many AR MDs shorten life expectancy, especially those of paediatric onset,[27, 29] and whilst this will not impact lifetime risk estimates, it substantially affects point prevalence (and to a lesser extent period prevalence) estimates. The only published prevalence estimate for nDNA associated MD, by Gorman and colleagues from an adult disease cohort, identified 2.9 affected individuals per 100,000, with half due to AR disease.[32] Paediatric MD estimates (of all MD, but predominantly comprising AR nDNA MD) have more often been reported as birth prevalence, ranging from 3.2 (1.7-5.4) to 15 (9.8-21) per 100,000.[10, 26–30] These data, all published prior to 2010, derive from regional laboratory data or clinical cohorts and do not include comprehensive genetic data. Although there is substantial variability in estimates, when considering the higher estimates – and incorporating adult prevalence (2.9 per 100,000) [32], advancements in diagnostic techniques, as well as non-conception and embryonic lethality rates [10] – the resulting figure begins to approach lifetime AR risk estimates from population-based data (25.8 per 100,000 individuals, 95% CI 18.2-32.9). This suggests that in favourable circumstances clinical identification of AR MD may be reasonably high, reflecting the tendency for such disorders to present as severe multisystem disease[2, 7], prompting early tertiary or specialist assessment. However, the wide range in estimates indicates other relevant contributing factors, which likely include the evolving biochemical diagnostic techniques and MD definition, the MD inclusion criteria required by the study, the population studied, as well as broader awareness and education throughout the medical fraternity impacting possible MD recognition, and access to care.[10, 26–29, 78] Incomplete penetrance of biallelic variants must also be considered, inviting consideration of polygenic risk, and unique to MDs, the possibility of bigenomic interplay on development of clinical disease. It is also important to note that the although assumptions underlying Hardy Weinberg equilibrium are rarely fulfilled,[48] it provides a useful paradigm for study. While our understanding remains incomplete, partly due to potential subclinical phenotypes, variable penetrance, ascertainment bias, and diagnostic limitations, these data provide valuable context for clinical sequencing results and genetic counselling. Combined with healthcare resource utilisation data,[11, 12] they may also help inform healthcare resource allocation and provision.

Estimating population-based MD risk associated with AD nuclear variants is a previously unaddressed challenge, complicated by the rarity of these disorders, the scarcity of pathogenic AD variants in healthy genetic reference populations, variable and age-dependent penetrance of the disorders and the prediction of novel variant heritability. Acknowledging these limitations, considering these findings can still offer insights into variant penetrance and expressivity.[79] Consistent with previous observations of pathogenic (non-MD) AD variants in healthy elderly,[80] we found dominant MD variants, considered pathogenic, in this healthy older population. Given the selection criteria for the MGRB cohort, it can be reasonably inferred that multisystem MD was absent in variant carriers.[50, 54, 80] If penetrance estimates from the literature are applied, the frequency of pathogenic AD variation in this healthy older cohort implies substantially greater population AD MD risk than disease ascertained clinically. This suggests that variants found incidentally may have lower penetrance than those identified in a clinical context, paralleling observations in mtDNA MD [71, 72] and other genetic diseases.[81, 82] Alternatively, expressivity may be insufficiently severe to be identified as MD, and various mechanisms potentially influence phenotype (for discussion, see [79]). This will have implications for clinical decision making, especially where identification of such variants is incidental. [80, 83] It invites further careful examination of clinical and genetic features in healthy carriers, to further our understanding of expressivity, penetrance and potential protective factors given the well elderly population evaluated in this study.[79, 80]

Diagnostic yield has improved incrementally with advancing knowledge and sequencing technology.[15, 18, 19, 41–45, 84, 85] However, variability in MD gene definition or gene set can have an important impact on identification and diagnosis of affected individuals, dependent on the scope, approach and availability of sequencing used in clinical practice.[15, 41–45, 84–92] Formation of an MD gene list revealed a core group of genes consistently associated with MD. However, there was substantial variability in the broader group of genes considered by each source, including by curated consensus clinical diagnostic tools, such as PanelApp.[14, 19, 21–25] These findings highlight the continually evolving understanding of MD genetic architecture and phenotypic variability,[14, 93, 94] the challenge of defining MD,[4] particularly as genes with a broader range of mitochondrial-associated functions are observed to effect mitochondrial phenotypes [14, 21] and the dynamic interchange between clinical diagnostic and research testing. In this study, gene set selection was observed to substantially impact MD lifetime risk estimates. In particular, the inclusion of genes more classically associated with metabolic disease notably increased risk estimates, underscoring the importance of a consensus definition of MD and MD-associated genes and consistent gene set curation for epidemiological comparisons and clinical practice.

It is important to note that relevant gene inclusions for clinical diagnostic purposes may differ from those considered relevant for pathophysiological or basic science research perspectives. For diagnostic purposes, when working from phenotype to genetic diagnosis, genes which produce a syndromic phenotype (or spectrum of syndromes) suggestive of MD should reasonably be included (i.e., a panel should incorporate MD genes and MD-mimic genes), such that clinical diagnostic yield is maximised. For research purposes, exploration of genes involved in mitochondrial function and pathophysiology may be of greater interest (depending on the research question), regardless of the phenotype or clinical syndrome and whether it is typical for MD. Equally, rationale for any gene exclusions is important. Genes that are clearly associated with mitochondrial dys/function, but with limited reports of disease to date, represent a conceptually different group from those where the mitochondrial association is unclear, or the mitochondrial association is definitive, but the phenotype is not classical for MD (such as Parkinson’s Disease or neurodegeneration with brain iron accumulation, NBIA). Extrapolating from the broadened phenotypic spectrum revealed by the advent of genetic diagnosis,[15, 93] it is possible non-classical MD phenotypes that clearly display mitochondrial dysfunction may in future be considered on the spectrum of phenotypic manifestations indicative of MDs. These shared pathophysiological mechanisms of disease will be especially important to consider as mitochondrial therapeutic avenues evolve.

The observed differences in gene sets by source also suggest locoregional differences in perspectives on MDs, which may reflect local population variation (e.g., founder effects or ethnically enriched variants), unique referral pathways, clinical practice expertise and research avenues. These factors should be considered in ongoing efforts to harmonise gene panels and build a conceptual understanding and definition of MDs applicable for clinical practice. Work toward consistent implementation of clearly-defined gene sets in practice (such as those by PanelApp and others [25, 95]) should seek to maximise diagnostic yield based on specialist evaluation of phenotype – thereby enabling optimal clinical care – without excessive analytic burden.

### Limitations

The approach to population risk estimation implemented for this study has inherent limitations, employing assumptions which are simplified and imperfect,[48] but is nevertheless of utility in considering and expanding MD epidemiology. This study has attempted to ensure comprehensive inclusion of MD genes and potentially deleterious variants. However, the genetics of MD are continuously evolving and some of the more recently identified MD-associated genes could not be included in the study (**Suppl1c_MDgenes_future**). Variant classification inherently introduces uncertainty, and whilst the approach to filtering and classifying variants was carefully considered to mitigate this, filtering by computational prediction was limited to embedded programs on the MGRB Vectis platform (SIFT and PolyPhen2), which are now superseded by more accurate and reliable tools. As such, classifications are continually evolving, and the evidence drawn upon is dynamic. In this context it is inevitable that classification will in some instances prove incorrect or change over time. This is anticipated to influence results in both directions, excluding pathogenic and including benign variants. Acknowledging the uncertainty inherent in this approach, inclusion of novel variants for more comprehensive risk estimation is justified by the observation that >50% of all variation is singleton and generally rare, even in very large genetic databases.[55] The relatively limited MGRB cohort size biases single-gene MD risk estimates, especially where only one or a small number of variants were identified (risk overestimation), and for those genes with no pathogenic alleles identified (risk underestimation). Consequently, summed MD risk across all genes may be more accurate than specific risk estimates for such individual genes. Variant detection was also limited to evaluating the presence of single nucleotide and small variants and could not detect larger structural variants and repeat expansions. There may be considerable disease burden caused by these other variant types, which are poorly resolved by short-read sequencing, and which may be more readily evaluated as long-read sequencing advances into mainstream practice.[96]

This study was performed in an Australian population cohort with predominantly European (non-Finnish) ancestry,[50] limiting generalisability outside of this population, especially given the observed variability of MD risk between populations. Population genetic databases disproportionately represent European ancestry with variation in other ethnic groups less well characterised,[97] resulting not only in potentially inadequate information for individual variant calls (impacting risk estimates), but also contributing to and entrenching health inequality.[98, 99] However, there were some clear strengths to using a cohort like this, as it was prospectively curated, had clear sample provenance, applied a consistent sequencing pipeline with clear quality control across all samples and implemented best practice guidelines, including for variant filtering.[50, 54] The cohort was well detailed genetically and clinically, and with the older age of participants, disease ought to have manifested by this later life stage, and if not manifest would likely be non-penetrant. The healthy sample also reasonably excludes multisystem disease in participants.[50] Therefore, although the sample is relatively small resulting in a wide error range, the estimates derived from this risk deplete cohort may reasonably be interpreted as minimum population risks and allow some inference to be drawn around penetrance for the modest number of AD MD variants identified. Finally, MD risk was estimated assuming chance coincidence of variants within a population; an assumption that would not be valid in populations with high consanguinity rates, as was previously observed in an Australian population subset,[10] although related individuals were removed from the MGRB cohort,[50] minimising any such effect.

## Conclusions

We observed a high rate of nDNA MD-gene associated variation in a healthy older Australian cohort, with an estimated AR MD lifetime risk of 25.8 (95% CI 18.7 to 32.9) per 100,000, corroborating MDs as one of the most common causes of heritable metabolic disease.[100] Around 1 in 500 individuals carried an AD MD variant, implying penetrance is lower than reported outside of the clinical context. Exact MD risk estimates vary substantially depending on the gene set considered, and a consensus approach to classifying MD genes is evolving. Contextualising sequencing data necessitates an understanding not only of overall MD risk but also of the population frequency of pathogenic variation and gene-specific MD risk, especially for the more common genetic forms of MD. Conceptually, population-based estimates of MD risk may offer an “upper bound” against which to evaluate clinical systems and optimise identification and diagnostic rates in clinical practice, ensuring appropriate pathways for access and resourcing of care. Through comparison with estimates derived from clinical cohort data, we infer that identification of nDNA MDs is variable. In optimal contexts, diagnostic rates across the lifespan may approach lifetime AR MD risk estimates, suggesting comprehensive ascertainment, whilst identification may be incomplete in many instances. Diagnostic rates will likely better approximate lifetime risk as comprehensive sequencing approaches become more widely available, facilitating appropriate planning of healthcare resources to benefit those affected.

## Supporting information

Supplementary 1_MD genes

Supplementary 2_AR variants

Supplementary 3_AR risk

Supplementary 4_AD variants

## Acknowledgements

E.C.W was supported by a New Zealand Neurological Foundation Chapman Fellowship (1955 CF).

R.L.D was supported by an NSW Health Early-Mid Career Fellowship.

C.M.S was supported by a National Health and Medical Research Council Practitioner Fellowship (APP1136800)

## Ethics

The ASPREE Biobank study was approved by the Alfred Hospital Human Research Ethics Committee, and the study was performed in accordance with ethical standards in the Declaration of Helsinki.

## Data Availability

The MGRB data used for this study was accessed via the open access Vectis platform, which is not currently active, although data is available on request at https://sgc.garvan.org.au/terms/mgrb/index.html.

## Supplementary Materials

Suppl1_MDgenes

Suppl2_ARvariants

Suppl3_ARrisk

Suppl4_ADvariants_table

## Author Contribution

E.W. conceptualisation, data curation, formal analysis, funding acquisition, investigation, methodology, validation, visualisation, writing – original draft and review & editing

S.R. data curation, resources and software, writing – review & editing

M.H. data curation, software, writing – review & editing

J.C. data curation, resources, software, writing – review & editing

C.Y. data curation, software, writing – review and editing

C.L. conceptualisation, investigation, methodology, supervision, validation, visualisation, writing – review & editing

S.K. resources, software, writing – review & editing

P.L. conceptualisation, resources, supervision, writing – review & editing

R.D. conceptualisation, investigation, methodology, administration, supervision, validation, visualisation, writing – review & editing

C.S. conceptualisation, funding acquisition, investigation, methodology, administration, resources, supervision, validation, writing – review & editing

## Use of AI statement

During preparation of this work, the authors used Google Gemma-3n-e4b AI Model on LM Studio to assist with editing. After using this tool, the authors reviewed and edited the content as needed, and take full responsibility for the content of the publication.

