## Supplementary 4_AD variants for "Population nuclear mitochondrial disease risk estimated from nuclear disease gene variants in a healthy older cohort"

**Table S1**. Mitochondrial disease (MD) variants identified in the Medical Genome Reference Bank (MGRB) cohort with potential autosomal dominant (AD) disease manifestations and estimated AD MD risk based on allele count and penetrance estimate.

| Gene | Variant (GRCh37) | Consequence | Inheritance | Allele count | Penetrance estimate (%) | Penetrance notes and references | MD risk  per 100,000 (95% CI) |
| --- | --- | --- | --- | --- | --- | --- | --- |
| *MFN2* | chr1-12069698-C-T | NM_014874.4(*MFN2*):c.2119C>T (p.Arg707Trp) | AR and AD | 1 | 50% | AD fully penetrant with age; however heterozygote parents reported asymptomatic. Of reported heterozygotes 9 of 17 symptoms or signs (53%; variable evaluation)(1-7) | 17.57  (0-129.58) |
| *OPA1* | chr3-193335054-T-A | NM_130837.3(*OPA1*):c.536T>A (p.Leu179Ter) | AD (inferred) | 1 | 80% | Truncating mutations approximately 80% penetrance in subtyped studies (depends on age, non-penetrance definition, careful evaluation)(8-12) | 28.12  (0.89-195.68) |
| *OPA1* | chr3-193353231-C-T | NM_130837.3(*OPA1*):c.868C>T (p.Arg290Ter) | AD | 1 | 80% | Truncating mutations approximately 80% penetrance in subtyped studies (depends on age, non-penetrance definition, careful evaluation)(8-12) | 28.12  (0.89-195.68) |
| *CYCS* | chr7-25163747-C-CTCTTTT | NM_018947.6(*CYCS*):c.-8-2_-8-1insAAAAGA, p.? | AD (inferred) | 1 | 100% | All variants to date with pedigrees suggest AD with full penetrance(13-17) | 35.15  (0.89-195.68) |
| *TWNK* | chr10-102748875-G-A | NM_021830.5(*TWNK*):c.908G>A (p.Arg303Gln) | AD | 1 | 60% | Minimum estimate from reported cases 60% (assumes all offspring inherited, age-appropriate careful exam), likely higher. NB single reported compound heterozygote with severe disease.(18-22) | 21.09  (0.89-195.68) |
| *IDH2* | chr15-90631934-C-T | NM_002168.4(*IDH2*):c.419G>A (p.Arg140Gln) | AD | 1 | 90% | High or full penetrance, excepting mosaicism(23, 24) | 31.63  (0.89-195.68) |
| *IDH2* | chr15-90633877-C-T | NM_002168.4(*IDH2*):c.208-1G>A (p.?) | AD | 1 | 90% | High or full penetrance, excepting mosaicism(23, 24) | 31.63  (0.89-195.68) |
| *SPG7* | chr16-89613064-AGGAGAGGCG-A | NM_003119.4(*SPG7*):c.1450-1_1457del | AR mostly, symptomatic heterozygotes | 3 | 5% | Small heterozygous risk alongside AR disease (controversial); age-related, variable expressivity (mild late cerebellar phenotype)(25-30) | 5.27  (0-129.58) |
| *POLRMT* | chr19-620488-C-G | NM_005035.4(*POLRMT*):c.2641-1G>C | AD (dominant negative) | 1 | 30% | Single report, no segregation, possible affected uncle. Use reduced penetrance estimate 30%(31) | 10.54  (0-129.58) |

All genes except *IDH2* were in the consensus list.
